# Cohort Profile of the Chilean COVID-19 Biorepository: a Multicentric initiative for multi-omics research on COVID-19 and LONG-COVID in a Latin American population

**DOI:** 10.1101/2023.12.20.23300304

**Authors:** Iskra A. Signore, Gerardo Donoso, Pamela Bocchieri, Eduardo A. Tobar-Calfucoy, Cristian E. Yáñez, Laura Carvajal-Silva, Andrea X. Silva, Carola Otth, Cappelli L. Claudio, Héctor Valenzuela Jorquera, Daniela Zapata-Contreras, Yolanda Espinosa-Parrilla, Paula Zúñiga Pacheco, Macarena Fuentes-Guajardo, Virginia A. Monardes-Ramírez, Pia Kochifas Velasquez, Christian A. Muñoz, Cristina Dorador, Jonathan García-Araya, Claudia P. Campillay-Véliz, Cesar Echeverria, Rodolfo Alejandro Santander, Leslie C. Cerpa, Matías F. Martínez, Luis Abel Quiñones, Eduardo Roberto Lamoza Galleguillos, Juan Saez Hidalgo, Estefanía Nova-Lamperti, Sergio Sanhueza, Annesi Giacaman, Gerardo Acosta-Jamett, Cristóbal Verdugo, Anita Plaza, Claudio Verdugo, Carolina Selman, Ricardo Alejandro Verdugo, Alicia Colombo

**Affiliations:** Department of Anatomic Pathology, Faculty of Medicine, University of Chile, Santiago, Chile; Service of Anatomic Pathology, University of Chile Clinical Hospital (HCUCH), Santiago, Chile; Human Genetics Program, Institute of Biomedical Science (ICBM), Faculty of Medicine, University of Chile, Santiago, Chile; AUSTRAL-omics, Vice Rector’s Office for Research, Development and Artistic Creation, Austral University of Chile, Valdivia, Chile; Institute of Environmental and Evolutionary Sciences, Faculty of Sciences, Austral University of Chile, Valdivia, Chile; Institute of Clinical Microbiology, Faculty of Medicine Austral University of Chile, Valdivia, Chile; School of Medicine, Magallanes University, Punta Arenas, Chile; Evolutionary and Medical Genomics of Magallanes (GEMMa), Assistance, Teaching and Research Center (CADI-UMAG), Magallanes University, Punta Arenas, Chile; Interuniversity Center on Healthy Aging, Chile; Department of Medical Technology, Faculty of Health Sciences, University of Tarapacá, Arica, Chile; Clinical Laboratory of the Technical Area of Molecular Biology, Salvador Hospital, Santiago, Chile; Department of Medical Technology, Faculty of Health Sciences, University of Antofagasta, Antofagasta, Chile; Laboratory of Microbial Complexity and Functional Ecology, Antofagasta Institute & Biotechnology Department, University of Antofagasta, Antofagasta, Chile; Laboratory of Molecular Virology, Faculty of Marine Sciences and Biological Resources, University of Antofagasta, Antofagasta, Chile; Laboratory of Molecular Biology, Nanomedicine and Genomics, Faculty of Medicine, University of Atacama, Copiapó, Chile; Emergency Public Assistance Hospital, Santiago, Chile; Emergency Medical Assistance Service (SAMU), Punta Arenas, Chile; Department of Basic and Clinical Oncology (DOBC), Faculty of Medicine, University of Chile, Santiago, Chile; Latin American Network for Implementation and Validation of Clinical Pharmacogenomics Guidelines (RELIVAF-CYTED), Madrid, Spain; Department of Pharmaceutical Sciences and Technology, School of Chemical and Pharmaceutical Sciences, University of Chile; Laboratory of Chemical Carcinogenesis and Pharmacogenetics (CQF), Department of Basic-Clinical Oncology (DOBC), Faculty of Medicine, University of Chile; Department of Computer Science, Faculty of Physical and Mathematical Sciences, University of Chile, Santiago, Chile; Clinical Biochemistry and Immunology Department, Faculty of Pharmacy, University of Concepción, Concepción, Chile; Center of Excellence in Translational Medicine, Faculty of Medicine, University of The Frontier, Temuco, Chile; Institute of Veterinary Preventive Medicine, Faculty of Veterinary Sciences, Austral University of Chile, Valdivia, Chile; Institute of Animal Pathology, Faculty of Veterinary Sciences, Austral University of Chile, Valdivia, Chile; Center for Surveillance and Evolution of Infectious Diseases, Austral University of Chile, Valdivia, Chile; Arturo López Pérez Foundation, Santiago, Chile; Institute of Interdisciplinary Research, University of Talca, Talca, Chile; School of Medicine, University of Talca, Talca, Chile

## Abstract

**Purpose:** Molecular mechanisms underlying COVID-19 susceptibility and severity are still poorly understood. The presence of genetic risk factors associated with ethnic background has been suggested, highlighting non-European ancestry as a risk factor for hospitalization in the United States. However, the representation of non-European populations in genomic case-control and cohort studies remains insufficient, and Latin American populations have been significantly understudied. Addressing this gap, we established The Chilean COVID-19 Biorepository, a multicentric endeavor comprising high-quality biological samples and associated data collected throughout Chile under stringent biobanking standards that ensure high quality, reproducibility, and interoperability.

**Participants:** The Chilean COVID-19 Biorepository was established by a network of nine nodes distributed in five macro-zones nationwide. The study enrolled adult participants living in Chile who had tested positive for SARS-CoV-2 infection and provided broad written informed consent. Blood samples were collected with EDTA and processed to store blood, plasma, buffy-coat, and DNA. Quality control measures, such as Standard Preanalytical Code (SPREC), incident reporting, DNA concentration, and absorbance ratio (260/280), were implemented to ensure the reliability and quality of the collected samples. Sociodemographic data, habits, clinical information, use of medications, and preexisting pathologies were registered. A weekly iterative workflow was implemented to ensure the quality and integrity of specimens and data.

**Findings to date:** Between October 2020 and February 2021, 2262 participants were recruited, pseudonymized, and categorized by disease severity into six categories, from asymptomatic to lethal. Notably, the Biorepository exhibited high compliance rates (>90%) across all quality control assessed items, reflecting high adherence to biobanking standards. A noteworthy feature of this cohort is the self-identification of 279 participants (12.3%) into thirteen different ethnic groups. Amerindian ancestry from genome-wide genetic data was 44.0%[SD15.5%] and increased to 61.2%[SD19.5%] when considering participants who identified as Native South Americans. As a data-contributor partner of the COVID-19 Host Genetics Initiative, the Chilean COVID-19 Biorepository has contributed to the publication of a second updated genome-wide association study, further enhancing our knowledge of the role of host genetics in susceptibility and severity to SARS-CoV-2.

**Future plans:** The Chilean COVID-19 Biorepository, under the leadership of Latin American researchers from a Latin American country, substantially adds to the integration of Latin American populations in the global collections landscape. Just as ocurred with the COVID-19 Host Genetics Initiative, we expect that this repository will attract global network collaborations for comparative studies on the effects of COVID-19 across diverse populations, including exploring potential genetic advantages or disadvantages in the context of SARS-CoV-2 infection. Researchers involved in establishing this biorepository are currently associated within a collaborative initiative known as COVID-19 Genomics Network (C19-GenoNet), aimed to accelerate the identification of genetic factors in both hosts and pathogens that influence the short and long-term outcomes of SARS-CoV-2 infection.

The broad informed consent utilized enables longitudinal cohort follow-up, thereby allowing for investigating the long-term consequences of SARS-CoV-2 infection, particularly concerning long-COVID. Thus, participants of this cohort were re-contacted to assess the development of long-COVID through a survey-based approach. The re-contact and recruitment procedures yielded a high response rate (82.11%), demonstrating strong participant engagement. In this case as well, this cohort has been leveraged by collaboration with the COVID-19 Host Genetics Initiative for the forthcoming publication of a genome-wide association study on long-COVID.

The concerted endeavors invested in this Chilean initiative have led to the establishment and consolidation of C19-GenoNet as both a research network and a biobanking network. A comprehensive catalog of the C19-GenoNet biobank network has been created and is accessible online at https://redcovid.uchile.cl/.

**STRENGHT AND LIMITATIONS OF THIS STUDY:**

- This study is one of the largest cohorts of COVID-19 patients with associated Biobank reported so far in Latin America.
- The study’s design and rigorous weekly monitoring ensured effective collection of high-quality simples and maximized the quality and completeness of data, with the ability to re-contact participants in case of problematic information.
- There were no control or reliable information about the time between the infection and the sampling, which may hamper the comparison of some parameters among cases due to transcriptional dynamics after SARS-CoV-2 infection.
- The study is based on a self-reported survey, which may represent a bias when analyzing specific clinical phenotypes.

## INTRODUCTION

The Coronavirus disease 19 (COVID-19) caused by the Severe Acute Respiratory Syndrome Coronavirus-2 (SARS-CoV-2) has led to a worldwide pandemic with severe consequences for public health. Since its first description in 2019, a broad spectrum of clinical manifestations [1,2], and several risk factors affecting disease severity (e.g. sex, age, comorbidities, previous health status and environmental conditions) have been reported [3].

Despite growing knowledge about the genetic and clinical factors associated with COVID-19 susceptibility and severity further research is needed. Several genome-wide association studies (GWAS) have shown an association between host genetic architecture and susceptibility to COVID-19 [4]. The COVID-19 Host Genetics Initiative reported a summary statistics meta-analysis in July 2021, finding 13 independent loci genome-wide significantly associated with SARS-CoV-2 infection or severe COVID-19 [5]. With its large and diverse sample size of 49,562 cases from 46 studies across 19 countries, this study significantly contributed to the identification of significant loci. The inclusion of European, admixed American, African, Middle Eastern, South Asian, and East Asian individuals [5] is particularly noteworthy since GWAS for COVID-19 have primarily been performed on populations of European ancestry.

The problem of low representation of ethnically diverse populations has been highlighted as it hampers translational research leading to inaccurate risk evaluations, inadequate public policies, and health inequalities [6,7]. Although Asian representation in Biorepositories and GWAS has notably increased in the last decade, African and Latin American populations are still underrepresented [8,9].

Nevertheless, the Centers for Disease Control and Prevention reported that in the United States, the COVID-19 hospitalization rate is between 4.6 and 5.3 times higher for Non-Hispanic Amerindians or Alaska Natives, Non-Hispanic black people, and Latinos when compared with Non-Hispanic white persons [10]. Also, Shelton and colleagues (2021) found that hospitalization rate for African Americans nearly doubled what would be expected based on the proportion of positive COVID-19 tests among the customers of a Direct-to-Consumer Testing company. Remarkably, non-European ancestry is a risk factor for hospitalization even after adjusting for sociodemographic and preexisting health conditions [11]. This difference cannot be explained by the two primary known genetic associations (ABO and 3p21.31 loci), suggesting other risk loci in the genome with ancestry-related allele frequency variations. Discovering the association between molecular profiles, epidemiological data, and the clinical outcomes in COVID-19 patients is critical to understand disease progression and improve diagnostic tests, drug development, and vaccine thus contributing to better public health strategies.

Biobank Networks swiftly supported global pandemic initiatives by providing the scientific community with samples, data, and collection and conservation standards and guidelines [12, 13]. For example, The United Kingdom Biobank and the pan-EU Biobanking and Biomolecular Resources Research Infrastructure generated large COVID-19 collections to study seroprevalence, its relationship with ethnicity, and comorbidities [11–14, 15]. However, there is still urgent to study non-European and US populations. Specifically, Latin American people with its diverse and mixed ancestry has been overlooked despite its potential to enrich genetic surveys. Therefore, it is essential to build COVID-19 biorepositories in Latin America that collect and conserve high-quality biological material and data. This paper reports the articulation of a network across Chile to recruit COVID-19 cases and the creation of a biorepository of samples and associated data collected under biobanking standards to ensure high quality, reproducibility, and interoperability.

## COHORT DESCRIPTION

### Study Design and Recruitment

The Chilean COVID-19 Biorepository, established by a network of nine nodes across five regions in Chile (Figure 1, left panel), received approval from the University of Chile’s Faculty of Medicine Ethics Committee and additional ratification from local Ethics Committees at each participating site (Ethics Committees of the University of Magallanes, Central Metropolitan Health Service, University of Tarapacá, and Scientific Ethical Committee at Valdivia Health Service). The study included individuals over 18 years of age who provided written broad informed consent (IC) to participate and to be re-contacted. Following international ethical recommendations [15], an adapted-for-biobanks broad informed consent was used in order to allow follow-up evaluations and capture and storage of samples and data for this and future studies on COVID-19 and other types of biomedical research. The management of donor recruitment, samples, and data was carried out in compliance with national laws [16,17], international ethical-legal regulations [18–20], and good biobank practices [21].

**Fig. 1.**
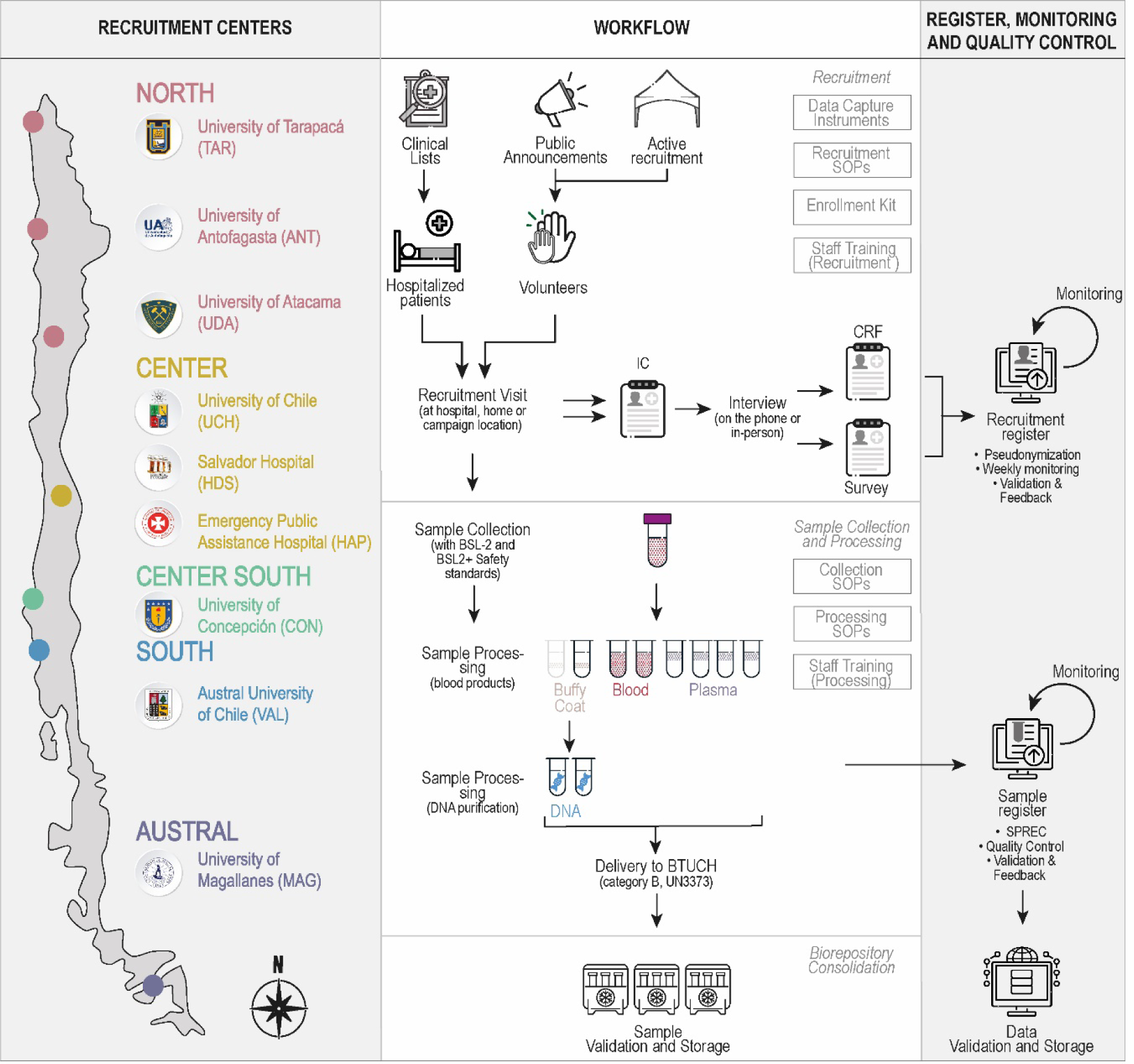
The Chilean COVID-19 Biorepository design and workflow. <u>Left panel.</u> Countrywide distribution of recruitment centers. Macrozones and the nine centers are located throughout the country, from the northern border to the southern region of the country. <u>Center Panel.</u> Workflow of the biorepository implementation showing Recruitment (top), Sample collection and Processing (middle) and consolidation (bottom). Processes performed in each phase are shown in boxes at the right corner of each subpanel. Volunteers joined the study through both active recruitment organized by centers and public announcement campaigns on social networks and the web. IC, CRF and Survey were obtained in person (at the centers or at the participant’s home), by telephone, or from the clinical records according to the requirements of each case. Processing of samples consisted of two phases: Blood samples were processed to obtain plasma (single spun) and buffy-coat, and then DNA from the latter. A complete set of samples included two tubes of blood (1 mL each), four of plasma (250 µL each), one of buffy-coat with 500 µL of RNA-later (Thermo Fisher Scientific), and two of DNA per donor (50-100 µL each). (see the text for further details). <u>Right Panel.</u> An online register system was implemented for monitoring and quality control. Data from CRF and Survey was collected in the Recruitment Register, while sample data were fed into the Sample Register. Registers were monitored and validated weekly to achieve the goals of the Chilean COVID-19 Biorepository (see the text for details).

Under the direct responsibility of the Biobank of Tissue and Fluids of the University of Chile (BTUCH), an intensive and centralized harmonization was developed to unify the (i) Standard Operating Procedures (SOPs) and Registers, (ii) sample coding system, and (iii) staff training for all the centers. A pilot phase was run to allow feedback and optimization of the first operational version of the standardized processes. All the documentation was centrally indexed in a digital online-shared Document Management Register for easy access. The document production and the staff training lasted one month. To minimize inconsistencies and errors in the required univocal assignment between codes and participants’ data and samples, an enrollment kit including all necessary documents (informed consent, case report form, epidemiological survey, and quality control records)and containers for every patient’s recruitment and sample collection was centrally pre-prepared with patient and sample code already assigned and affixed, validated, and sent to each center by the BTUCH. The pilot phase took place in September 2020, while official sample and data capture started in October 2020 and continued until May 2021.

Patients joined the study in two ways: invitations based on lists of hospitalized patients and recruitment of volunteers (Figure 1, top center panel). Hospitalized adult patients admitted during the study were included after a positive result on a reverse-transcriptase coupled to quantitative polymerase chain reaction assay (qRT-PCR) test on a nasopharyngeal swab. Only laboratory-confirmed cases with test results available through electronic medical records were included. Patients admitted for other reasons and circumstantially found COVID-19-positive were excluded. For non-hospitalized patients, confirmed infection of SARS-CoV-2, either by laboratory testing (SARS-CoV-2 qRT-PCR test, SARS-CoV-2 Rapid Antigen Test, Antibody Tests for COVID-19) or clinical diagnosis, was required. Self-report of suspected disease was not included in the study.

Each center maintained an online recruitment registry for weekly follow-up to evaluate and improve recruitment dynamics. IC and subsequent Case Report Form (CRF) or Survey (Figure 1, top center panel) were obtained by specifically trained personnel to collect contact information, demographic variables, COVID-19 symptomatology, ethnicity, ancestry, personal medical history, comorbidities, medication, and other relevant clinical data (Figure 2 and Supplementary Tables 1 and 2). The REDCap platform (https://www.project-redcap.org/) was used for data management. Case and sample data were digitized, pseudonymized, and reviewed locally before being sent to the central biorepository for validation and logging into the central collection database’s Recruitment Register. Weekly monitoring of this register provided detailed feedback for the continual improvement process (Figure 1, top right panel) allowing the development tailored strategies to address each center’s unique challenges.

**Fig 2.**
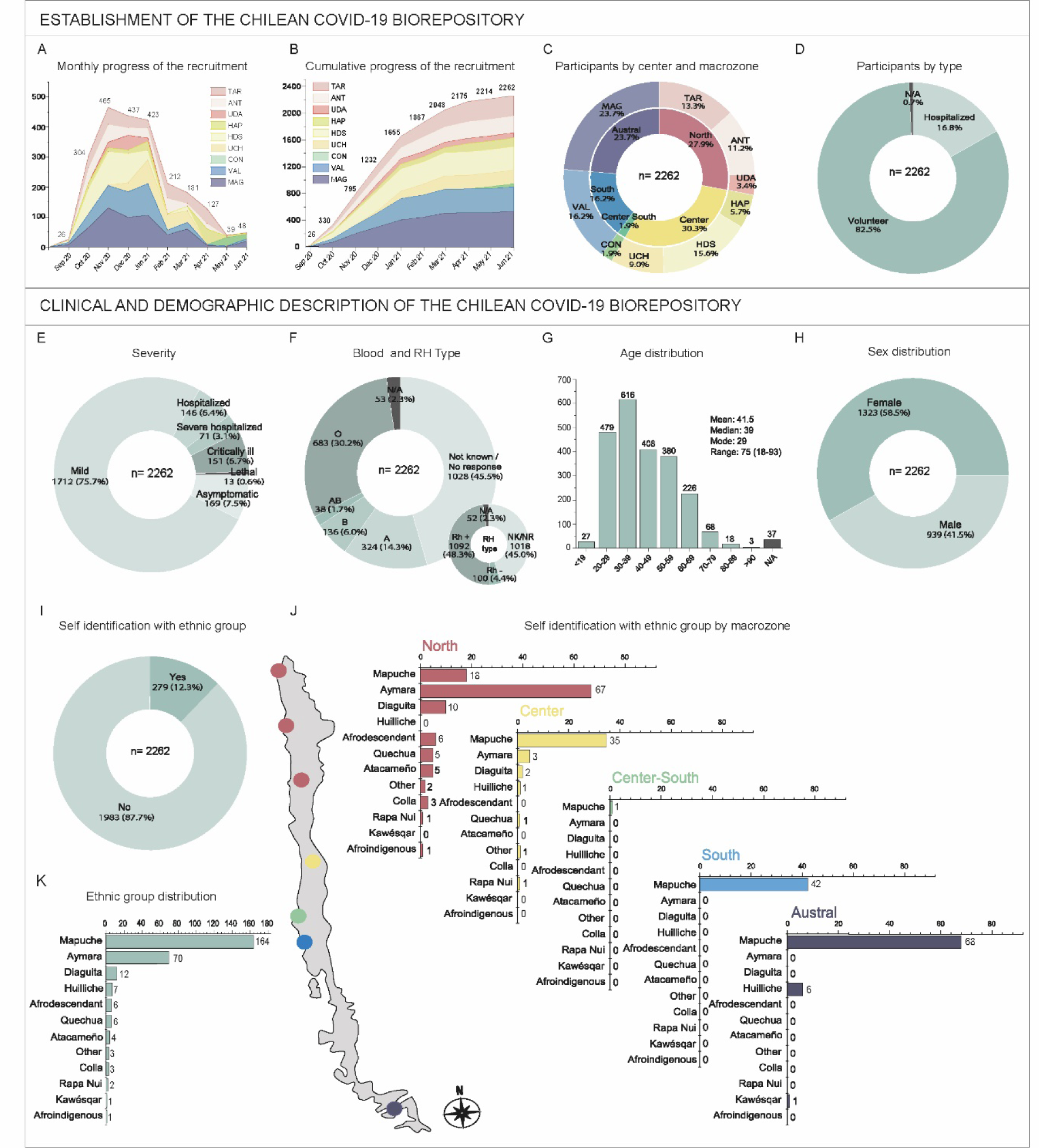
Description of the dynamics of recruitment and donors’ information of the Chilean COVID-19 Biorepository. <u>Top panel</u>. Establishment of the Chilean COVID-19 Biorepository (A-B) Recruitment progress. Cases registered in September 2020 correspond to the pilot (see text), while those that appear in June 2021 correspond to donors whose date of capture was corrected *a posteriori*, presented errors, inconsistencies, or incomplete data. (C) Concentric graph showing in the inner ring the distribution of participants captured by macrozones in correspondence with related centers shown in the external ring. (D) Proportion of participants entered through volunteer campaigns or hospitalization. The cases indicated as N/A (n=15) correspond to those data that presented errors, inconsistencies, or were absent. <u>Bottom panel.</u> Clinical and demographic description of the Biorepository. (E) Proportion of patients according to severity. (F) Self-reported blood and RH-type. NK: not known NR: no response. (G) Histogram depicting the age structure of the biorepository cohort. The descriptive statistics parameters mean, median, mode, and range are shown, with the youngest patient being 18 years old and the oldest being 93 years old. The 44 cases labeled N/A correspond to inconsistent, erroneous or missing data, as in the previous panel. (H) Proportion of female and male participants in the biorepository showing that most of them were women. (I) 386 participants self-identified as belonging to an ethnic group. Cases labeled N/A correspond to inconsistent, erroneous or missing data. The frequencies of each group at the whole biorepository level are shown in K, sorted from highest to lowest. The order obtained is maintained in J to show the distribution of these ethnicity groups within each macrozone.

### Sample collection and processing

As depicted in Figure 1 (middle panel), a volume of 6 mL of blood sample was collected in a tube with EDTA anticoagulant for each patient. In 55 cases, the sample was collected by buccal swabbing as the patients had poor venous access. At the corresponding center, blood samples were processed using a two-step process performed by dedicated personnel whose high quality and consistency skills were monitored and, when necessary, improved, and re-trained (Figure 1, middle panel). After testing its equivalence through a comparative analysis of quality and performance, DNA was purified using either an EZNA blood DNA min kit (Omega Bio-Tek) or QIAamp DNA mini-Blood (Qiagen, Inc.). At the end of the process, a complete set of samples comprised 9 tubes, Buccal swabbing samples were entirely used to purify DNA, being the only stored biospecimen in these cases. Samples were collected and processed in compliance with biosafety guidelines for levels 2+ and 2 (BSL-2 and BSL2+) [22].

The Standard Preanalytical Code (SPREC) was applied for quality control (QC) of the procedure, which involved recording the collection time, the start time of centrifugation, and the commencement of long-term storage [24]. Additionally, QC of the sample included incident reporting for blood and blood product samples (hemolysis, jaundice, lipaemia, and clotting). The quality of DNA samples was monitored by measuring volume, concentration (ng/µL), and the absorbance ratio A^260/280^. Sample data were reviewed at each center and logged to the Sample Register at the central collection database for validation and feedback (Figure 1, center right). Finally, samples were delivered to the BTUCH (−33.4194 −70.6529) using a triple packaging system complying with UN3373 standards [23] for endmost validation and long-term storage (Figure 1, bottom middle panel).

### DNA genotyping and estimation of ancestry

Genome-wide genotypes were generated by hybridization to Global Survey Arrays v3 (Illumina Inc.). Individuals and Single Nucleotide Polymorphisms (SNPs) with more than 5% missing data were removed. In addition, SNPs with p-values < 10E-6 for Hardy-Weinberg disequilibrium and MAF<0.01 were also removed. Genotypes were merged with a reference data set of 40 European (CEU), 40 African (AFR), and 40 east Asian (EAS) individuals from the 1000 Genomes Project [24], 40 individuals of Aymara ancestry (Personal communication, Andrés Moreno-Estrada) and 40 individuals of Mapuche ancestry [25,26]. Global ancestry was estimated using ADMIXTURE v1.3.0 using supervised clustering [27]. The best number of estimated subpopulations (k-value) was chosen to minimize cross-validation error.

### Establishment of the Chilean COVID-19 Biorepository

By centralizing the coordination of the network, 2262 participants were enrolled in 8 months ensuring a high standard of sample and data quality. The recruitment was preceded by a pilot performed at Salvador Hospital in September 2020. Figures 2A and 2B show the recruitment progress, revealing that although each center had a different dynamic, the most intense recruitment period was between October 2020 and February 2021, with a peak of 465 participants in November 2021. Except for the University of Concepción (CON; Figure 2A and 2B in green), all centers initiated and maintained recruitment efforts consistently throughout the project’s 8-month recruitment phase. Although centers are differently distributed on the national territory, the contribution of the macrozones was remarkably even (Figure 2C) with the sole exception of the Center South macrozone (contribution 1.9%). This is noteworthy considering the high centralization of the country in terms of population, accessibility, and general resources. Regarding the routes of admission to the biorepository, 1866 donors were volunteers while 381 were hospitalized patients (Figure 2D). The remaining 15 cases have missing or inconsistent information on this point (0.7%, indicated as N/A in Figure 2D)

### Demographic and clinical description of the Chilean COVID-19 Biorepository

Participants were categorized by disease severity into six categories: asymptomatic (169), mild (1712), hospitalized (146, not requiring oxygen support), severe hospitalized (71, requiring oxygen support), critically ill (151, requiring respiratory support with mechanical ventilation such as continuous (CPAP) or bilevel (BiPAP) positive airway pressure or Optiflow/very high flow Positive End Expiratory Pressure Oxygen (PEEP), and lethal (13) (Figure 2E).

Using survey and CRF, we collected data on participants’ clinical information (blood group, body mass index (BMI), chronic and preexisting pathologies, use of medications), sociodemographic characteristics (sex, age, weight, height, level of schooling, private/public healthcare, self-identification with ethnic groups, ancestry), and habits (tobacco, alcohol, and drugs consumption). Figure 2 shows some of these data. Self-reported blood and Rh type showed a large proportion of individuals who did not know their ABO and Rh blood group. Neverthess, the most common types in the biorepository are O and Rh+ (Figure 2F). The age distribution and its descriptive statistics are shown in Figure 2G but most participants were between 30 and 39 years old. Females account for 58.5% of the biorepository (Figure 2H), A noteworthy feature is the presence of 279 self-identifying members of an ethnic group (12.3% of the repository) (Figure 2I). Members of eight of the ten officially recognized native peoples in Chile were present, along with Huilliche, Afro-descendants, and Afro-Indigenous (Figure 2K). Figure 2J shows the distribution of ethnic groups by macrozones and some geographic correlation between the two can be appreciated.

### Patient and public involvement

Participants were not involved in the design stage. As the recruitment commenced, we incorporated their input and needs, modifying our methodology accordingly. For instance, we arranged home visits to ease participation and minimize discomfort and costs associated with obtaining informed consent, survey completion, and sample collection. When visits to the recruitment center were preferred, we scheduled them to coincide with participants’ pre-existing medical appointments at hospitals and medical centers where our recruitment centers were located. Taking participant suggestions onboard, we implemented an online survey option to facilitate participation remotely, forwarding the link via email or chat. To increase reciprocity and trust, we asked participants about their data return preferences, resulting in the idea of providing personalized reports detailing the findings of their ancestry analysis as a way to return research data to participants.

## FINDINGS TO DATE

### Description of samples collection

The Biorepository is physically located at BTUCH within the Clinical Hospital of the University of Chile, in Santiago, (coordinates −33.4199, −70.6531). For 2057 out of 2262 (88.9%), we collected a complete set of biospecimens while at least one sample was collected from 205 donors (8.9%). Notably, no sample was obtained in only 52 cases (2.2%) (Figure 3A). The average effectiveness of centers in obtaining the entire set of biospecimens per patient was 91.7% (minimum value 78.0%), while the average of obtaining no sample per patient was 1.6%, (maximum value 4.7%). The high capture effectiveness show that our multicenter design achieved good national ongoing coordination (Figure 3B and 3D).

**Fig 3.**
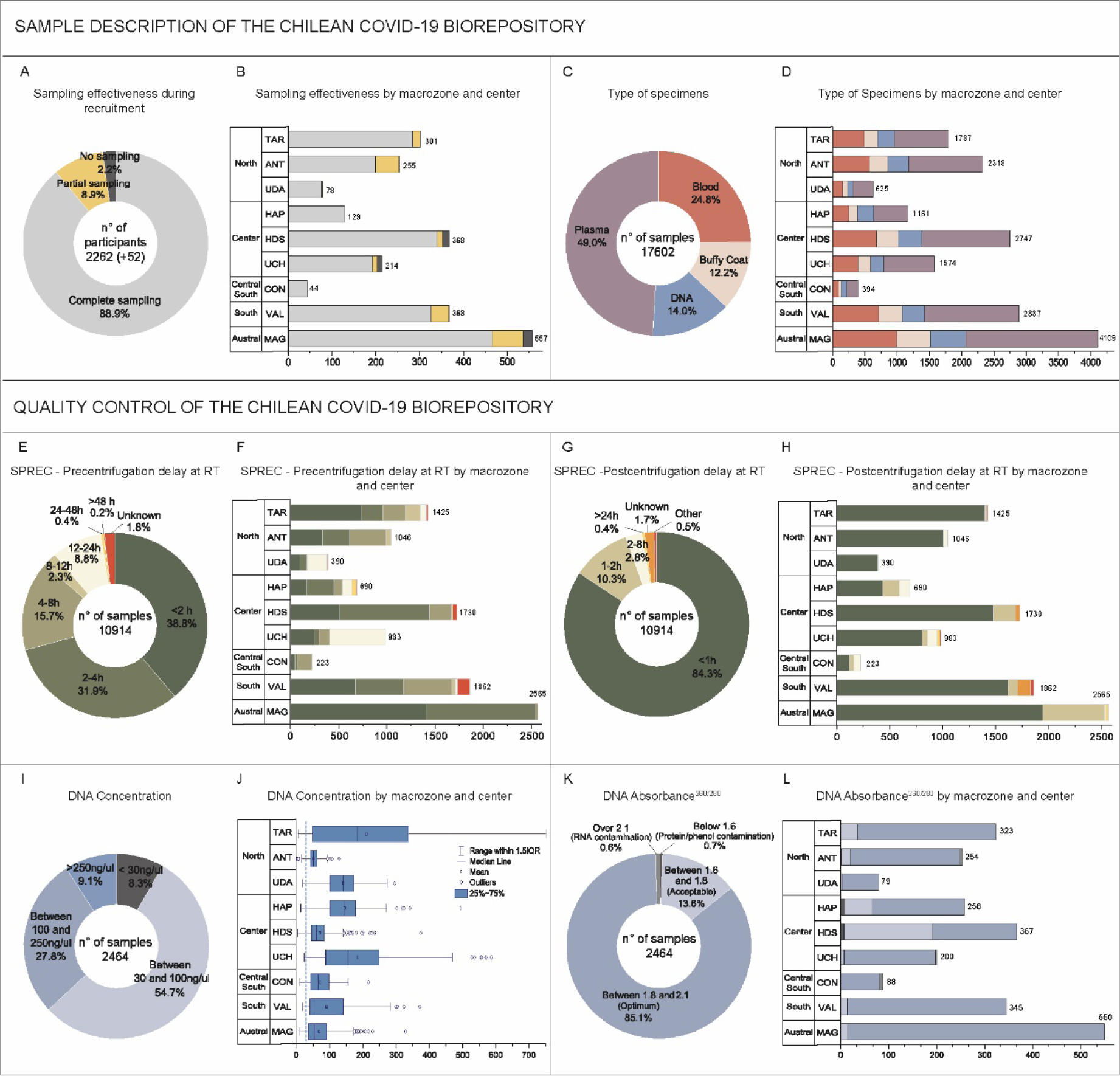
Description of the type and quality of the biological samples of the Chilean COVID-19 Biorepository. <u>Top panel.</u> Sample description. (A-B) Performance of the centers concerning the collection of donor samples. Only 52 participants were left out of the biorepository due to sample collection failure (in black). For the remaining 2262 cases, a final set of 9 or 8 sample tubes was considered a complete sampling since it includes all necessary specimen types (see Figure 1). Donors with a final set of between 1 and 7 specimens were considered partial sampling. (C-D) Distribution of specimens by type in the whole repository and by macrozone and center of the whole repository. <u>Bottom panel.</u> Quality Control. (E-H) Standard preanalytical code (SPREC) for pre (E-F) and post (G-H) centrifugation delay at Room Temperature. The compliance with the required standard is depicted in all shades of green, while values below the acceptable standard are in yellow, orange, and red and correspond to values larger than 24hrs for both pre- and post-centrifugation as well as incompletely recorded cases (labeled as unknown) and those with other notes that do not correspond to any other SPREC code (labeled as other). (I-L) Quality for DNA in terms of concentration (I-J) and purity (K-L). Samples that meet the standard established for the biorepository are shown in shades of blue, while non-compliant samples are in gray. In (J), the dotted line corresponds to the minimum concentration standard of 30 ng/µl.

The total number of biospecimens is 17602, distributed as follows (cryotubes/different samples: Blood (4357/2198), Buffy-coat (2150/2150), DNA (2464/2186) and Plasma (8631/ 2159) (Figure 3C). Incidents were reported only in 82 cases (3.6%) and were hemolyzed (n=36), lipemic (n=24), icteric (n=10), and clotted (n=2) samples, plus 10 tubes in which the minimum collection was not achieved.

We performed QC of our preanalytical phase according to SPREC, confirming the proper handling of the samples. In this case, our efficiency in recording the information necessary to determine SPREC codes was lower: out of 17602 total samples, we have 10914 quality records of which, only 266 (2.4%) and 282 (2.6%) did not meet pre- and post-centrifugation delay requirements, respectively (Figure 3E-3G sum of yellow, orange, and red). For pre-centrifugation delay, only 3 centers performed under the 95,0% of complying (Figure 3F; 92.8% HAP, 93.0% VAL, and 94.9% UDA), while for post-centrifugation there was only one (Figure 3H: 91.7% VAL).

The minimum quality standard we set for the DNA concentration was 30 ng/µl, suitable for most subsequent technical needs, including sequencing studies. Considering the whole repository, 2259 samples out of 2464 fulfilled this requirement reaching compliance of 91.7% (Figure 3I, all shades of blue) with an average DNA concentration of 109,1ng/µL. Remarkably, 37.0% of the DNA samples (911 biospecimens) had a concentration above 100ng/µL (Figure 3E). Only 3 out of 9 centers had an average compliance rate below 90%

(60,5% MAG, 77.6% TAR, and 88.8% VAL) for this standard of 30ng/mL but all centers achieved a mean concentration well above of this limit as shown in Figure 3J. The analysis of the absorbance ratio 260/280 of DNA samples revealed that only 32 samples (1.3%) were contaminated either with protein, phenol, RNA, or other contaminants (Figure 3K, in shadows of grey). Most DNA samples (2098 specimens) showed optimal quality with absorbance between 1.8 and 2.1, while 334 had acceptable quality with absorbance between 1.6 and 1.8 (Figure 3K, all shades of blue). The performance of the different centers was once more even, being 93.2% of uncontaminated samples the lowest level of compliance regarding DNA absorbance values (Figure 3L).

### Description of symptoms and comorbidities

Participants of the Chilean COVID-19 Biorepository reported 34 previously described symptoms. The list of symptoms included in the survey was compiled from literature, the COVID-19 Participant Experience-COPE [28] and a preliminary version of the LMIC LPS-Covid-19-Questionnaire generated by a working group hosted by Wellcome and the International Hundred Cohort Consortium-IHCC and made available to this study by personal communication with Dr. Teri Manolio. Symptoms ranged in frequency from 3.0% to 68.1% for skin discoloration and fatigue, respectively. Table 1 shows the five most common symptoms by severity level. Expectedly, respiratory symptoms like dyspnea (both at exertion and rest) increase in more severe disease forms leading to hospitalization. Full list of symptoms and detailed frequencies are available in Supplementary Table 1. Also, 26 comorbidities were reported, and obesity, hypertension, diabetes, high cholesterol, and other chronic diseases was found in all conditions. The five most common comorbidities by severity are also shown in Table 1, and the full list is available as Supplementary Table 2.

**Table 1.** Prevalence of the five most common symptoms and comorbidities reported in Survey and CRF in the whole biorepository and by severity level.

| SYMPTOMS |  |  |  |  |  |
| --- | --- | --- | --- | --- | --- |
| Whole Biorepository |  | Mild |  | Hospitalized |  |
| Fatigue | 1541 (68.1%) | Fatigue | 1355 (79.1%) | Dyspnea on exertion<sup>s</sup> | 98 (67.1%) |
| Headache | 1521 (67.2%) | Headache | 1326 (77.5%) | Pneumonia | 88 (60.3%) |
| Myalgia | 1365 (60.3%) | Anosmia | 1193 (69.7%) | Fatigue | 84 (57.5%) |
| Decay | 1322 (58.4%) | Decay | 1160 (67.8%) | Headache | 82 (56.2%) |
| Anosmia | 1305 (57.7%) | Myalgia | 1157 (67.6%) | Fever | 80 (54.8%) |
| Severe hospitalized |  | Critically ill |  | Lethal |  |
| Dyspnea on exertion<sup>s</sup> | 58 (81.7%) | Dyspnea on exertion<sup>s</sup> | 107 (70.9%) | Dyspnea on exertion<sup>s</sup> | 7 (53.8%) |
| Fatigue | 57 (80.3%) | Myalgia | 83 (55.0%) | Fever | 5 (38.5%) |
| Pneumonia | 56 (78.9%) | Fever | 81 (53.6%) | Persistent dry cough | 3 (23.1%) |
| Difficulty breathing at rest | 51 (71.8%) | Other symptoms | 81 (53.6%) | New productive cough | 2 (15.4%) |
| Decay | 51 (71.8%) | Headache | 65 (43.0%) | Diarrhea | 2 (15.4%) |
| COMORBIDITIES |  |  |  |  |  |
| Whole Biorepository |  | Asymptomatic |  | Mild |  |
| Other chronic diseases | 719 (31.8%) | Obesity | 55 (32.5%) | Other chronic diseases | 571 (33.4%) |
| Obesity | 691 (30.5%) | Other chronic diseases | 38 (22.5%) | Obesity | 529 (30.9%) |
| Hypertension | 394 (17.4%) | Hypertension | 27 (16.0%) | Hypertension | 212 (12.4%) |
| Mental health problems | 233 (10.3%) | Diabetes | 21 (12.4%) | Mental health problems | 185 (10.8%) |
| High cholesterol | 232 (10.3%) | Mental health problems | 17 (10.1%) | High cholesterol | 161 (9.4%) |
| Hospitalized |  | Severe hospitalized |  | Critically ill |  |
| Other chronic diseases | 51 (34.9%) | Obesity | 32 (45.1%) | Hypertension | 72 (47.7%) |
| Hypertension | 49 (33.6%) | Hypertension | 26 (36.6%) | Diabetes | 35 (23.2%) |
| Obesity | 39 (26.7%) | Other chronic diseases | 24 (33.8%) | Obesity | 35 (23.2%) |
| Diabetes | 28 (19.2%) | Diabetes | 23 (32.4%) | Other chronic diseases | 31 (20.5%) |
| High cholesterol | 25 (17.1%) | High cholesterol | 12 (16.9%) | High cholesterol | 17 (11.3%) |
| Lethal |  |  |  |  |  |
| Hypertension | 8 (61.5%) |  |  |  |  |
| Other chronic diseases | 4 (30.8%) |  |  |  |  |
| Cardiac problems | 3 (23.1%) |  |  |  |  |
| Diabetes | 2 (15.4%) |  |  |  |  |
| Coronary atherosclerosis | 2 (15.4%) |  |  |  |  |

### Description of ethnic composition and ancestry

At the time of writing this paper, validated and quality-controlled ancestry analysis were completed for 2099 participants. The remaining 163 (7.2%) are cases in which the sequencing run failed, or the DNA concentration or quality were inadequate and, thus, were not included in the run. For these cases, participants were re-contacted new samples have been taken from those who agreed, and the DNA is being sent for a new analysis. Thus, the ancestry of the whole cohort was determined using genome-wide genotyping data from the 92.8% of the samples, resulting in an average distribution of 2.0% African [SD 3.6%], 1.6% Asian [SD 4.2%], 52.4% European [SD 15.2%], and 44.0% Amerindian [SD 15.5%].The latter ancestry can be decomposed into 13.4%[SD 17.2%] of a northern component that is more closely related to Aymara and Quechua ethnicities and 30.6%[SD 15.0%] of a southern component more closely related to Mapuche ethnicity. The Amerindian component was 61.2%[SD 19.5%] in individuals who self-identified with any Native American ethnic group. Details on ancestry by self-declared ethnicity can be found in Table 2.

**Table 2.** Average (SD) of ancestry by self-declared ethnicity.

| Self-reported Ethnicity | N | African (AFR) | Asian (EAS) | European (EUR) | Northern Andes | Southern Chile | Amerindian<sup>4</sup> |
| --- | --- | --- | --- | --- | --- | --- | --- |
| African | 6 | 24.8 (32.8) | 1.0 (1.8) | 36.2 (16.7) | 23.5 (14.1) | 14.5 (8.7) | 37.9 (17.3) |
| Native American | 252 | 1.4 (1.8) | 0.9 (1.4) | 36.5 (18.5) | 24.6 (32.5) | 36.6 (24.1) | 61.2 (19.5) |
| Andes North<sup>2</sup> | 79 | 1.9 (2.6) | 0.9 (2.0) | 22.9 (20.1) | 64.4 (31.0) | 10.0 (9.5) | 74.4 (22.4) |
| Chile South<sup>3</sup> | 162 | 0.9 (0.9) | 0.8 (1.0) | 42.4 (14.2) | 5.6 (6.1) | 50.3 (17.7) | 55.9 (14.8) |
| Diaguita | 11 | 4.2 (1.3) | 1.1 (1.0) | 48.8 (6.5) | 19.5 (4.2) | 26.5 (5.3) | 45.9 (6.0) |
| Rapa Nui | 2 | 7.7 (8.4) | 1.0 (1.1) | 41.3 (29.9) | 28.5 (28.3) | 21.5 (7.9) | 50.0 (20.4) |
| Other | 3 | 2.3 (0.6) | 1.7 (2.1) | 44.3 (22.7) | 23.8 (28.5) | 28.0 (10.4) | 51.7 (20.9) |
| None declared | 1836 | 2.0 (3.1) | 1.7 (4.5) | 54.7 (13.2) | 11.9 (13.2) | 29.8 (13.1) | 41.7 (13.2) |
| Whole Cohort | 2099 | 2.0 (3.6) | 1.6 (4.2) | 52.4 (15.2) | 13.4 (17.2) | 30.6 (15.0) | 44.0 (15.5) |

<sup>1</sup>Ancestry was estimated using ADMIXTURE on genotypes generated by the GSA v3 microarray
<sup>2</sup>Self-identified as Atacameño, Aymara, Quechua or Colla
<sup>3</sup>Self-identified as Huilliche, Mapuche, or Kawésqar
<sup>4</sup>Amerindian ancestry for each participant was estimated by adding the Northern Andes and Southern Chile ancestries

## STRENGHT AND LIMITATIONS

The outbreak of the COVID-19 pandemic impels the scientific community to understand the variable manifestations and consequences of the infection at the individual and population levels. Here we describe one of the largest prospective cohorts of COVID-19 patients with associated Biobank reported so far in Latin America together with the BRACOVID cohort (5233 participants) [29]. Since the population of Brazil is almost 11 times that of Chile, the Chilean COVID-19 Biorepository is a remarkable effort. The design of the cohort was addressed toward the identification of genetic variation in the Chilean population associated with the severity and clinical evolution of COVID-19. The cohort comprised individuals from north to south of Chile and the ancestry of our cohort is highly admixed, closely resembling the genetic profile of the general Chilean population [30,31]. Notably, in the GWAS studies conducted by the COVID-19 Host Genetics initiative and by 23andMe, the percentage of European ancestry ranged from 71-78% [5] to 80.3% [32], respectively. A similar percentage of 70% was reported in another study among participants of the Million Veteran Program [33]. These results highlight the importance of Latin American biorepositories like this, whose European composition ranges from 23.4% to 55.8%, depending on the self-identified ethnicity (Table 2).

Three years after the COVID-19 pandemic onset, predicting the severity of the disease remains challenging. Age and sex are confirmed risk factors, with higher disease severity described for older male individuals [34]. Preexisting conditions such as hypertension and diabetes mellitus have been described to increase the severity of COVID-19, but the specific association between comorbidities and the prognosis remains unclear [35,36]. Regarding the blood group, the impact on clinical outcome is unclear, but the influence on susceptibility to infection appears to be very likely (reviewed in [37]). Figures 2 and 3 show that our biorepository can be relevant to study these aspects.

One of the strengths of this project and its major challenge was the countrywide multicenter structure. The design aimed to obtain consistent quality of samples and data while promoting heterogeneity and reducing common biases occurring in multicenter recruitment, for example, due to the presence of large urban centers. First, we made a considerable effort to standardize procedures by centralizing critical processes such as staff training, digital document management, and the preparation of the recruitment kit. Secondly, a strategy of continuous and rigorous monitoring was adopted based on weekly automatic data-check and virtual meetings. This approach allowed us to readily identify problems, inconsistencies, and incompleteness on the spot and apply remedial strategies promptly. For example, matrix for DNA extraction was changed, favoring purification from buffy-coat over purification from blood. Also, the need to reinforce the training of the personnel in charge of DNA purification was detected very early, and the competence for this specific task was readily improved. Our experience shows that a high level of standardization of biobanking processes is necessary to obtain good results, but it is not sufficient alone as it cannot replace nor compensate for the expertise of the personnel in charge.

Our design resulted in high effectiveness in collecting blood samples (Figure 3A) and high-quality blood products and good DNA concentrations (Figure 3I). The SPREC QCs also supported this result, but improving the complete capture of this information is a point to be addressed in the future.

Weekly monitoring during recruitment maximized the quality and completeness of data by allowing re-contacting participants in the case of problematic information in the CRF or Survey. This strategy resulted in a very low number of inconsistencies, absences, or errors in data collection (indicated as N/A in Figures 2 and 3), which affected a maximum of 53 cases, corresponding to less than 2.5% of the participants. Our results show that our strategies worked, and that the proposed aims regarding the composition and quality of the Chilean COVID-19 Biorepository were fully achieved.

Implementation of this biorepository can have an impact both nationally and internationally across multiple public health perspectives. We have already highlighted that lack of diversity and underrepresentation of Latin Americans hinders translational medicine and medical-clinical research and how this problem need to be addressed to design public health policies accurately-targeted to their intended populations. [38–41]. Undoubtedly, the Chilean COVID-19 Biorepository is an important contribution in this sense. Also, the Chilean population is highly heterogeneous, and its diversity is not uniformly distributed throughout the country in association with its geography [31]. The multicenter design organized by macrozones was chosen especially considering a possible differential susceptibility of different populations to COVID-19. However, even if the collection comprised 2262 cases, particular ancestry groups such as Aymara or Mapuche constitute only a small subset of the sample. Similarly, most of the recruited cases suffered from mild COVID-19 so that very severe COVID-19 are less represented. Also, the study is based on a self-reported survey which may represent a bias when analyzing specific clinical phenotypes.

From the ethical point of view, the use and acceptance of broad informed consent are established but rather recent in the country. Although in our legislation the ethical authorization of an accredited committee allows studies to be carried out in any region of our country [16], a new revision by the Ethics Committee at each recruitment center was necessary. This delayed the project and impacted on the number of participants that could be recruited with the approved budget. It is important to consider local particularities, but it is also essential to advance in national policies that safeguard the rights and welfare of participants while allowing faster implementation of scientific studies, especially when they can help orient public policies in times of pandemics [15,42].

Motivating people to participate in scientific studies is always a challenge in which biobanks play a positive critical role. Unfortunately, there is no national policy in Chile for the creation and operation of biobanks, leading to a lack of direct or competitive funding so that is difficult to maintain the few existing biobanks, particularly outside the nation’s capital. This over-centralization problem threatens valuable biological materials that need to be shipped to Santiago for long-term storage. The COVID-19 pandemic demonstrated the importance of biobanks to support the basic-clinical research needed to respond to emergencies. Chile lacks a population biobank with international standards and national representation, but the collaboration of the institutions involved in this project succeeded in creating this Chilean COVID-19 Biorepository.

## ETHICS APPROVAL

The study was conducted under ethical and scientific standards and was approved by the relevant institutional ethical review boards. Further detailed information can be found in the text.

## CONSENT TO PARTICIPATE

Informed consent was obtained from all individual participants included in the study.

## COLLABORATION

Researchers from any location can submit applications for accessing the Chilean COVID-19 Biorepository. Data and samples may be made available upon request and after ethical and scientific evaluation and approval to ensure compliance with national laws and relevant policies of the biobanks, institutions and researchers involved in the creation of biorepository and members of the C19-GenoNet biobank network. Individual-level ancestry estimates for European, African, Asian, and Amerindian origins are available along with demographic and clinically relevant variables.

## AUTHORS CONTRIBUTIONS

The authors have contributed to this work as follows. Conceptualization: AXS, YEP, LAQ, RAV, AC; Methodology: GD, PB, LAB, AC. Software: JSH; Formal analysis: IAS; Investigation, sample collection and processing: ETC, CY, LCS, CCL, HVJ, DZC, PZP, VAMR, PKV, CAM, JGA, CPCV, CE, RAS, LCC, MFM, ERLG, ENL, SS, AG, GAJ, CbV, CV, AP; Resources: GD, PB, MFG, CD, LAQ, ENL, AC; Data curation: IAS, GD, ETC, CY, MFG, LCC, JSH, RAV; Writing the original draft: IAS, AC; Reviewing and editing the draft: IAS, AXS, DZC, YEP, MFG, LAQ, ENL, RAV, AC; Visualization of data: IAS, JSH; Supervision: GD, PB, CCL, AXS, DZC, VAMR, CAM, CD, MFM, CS, RAV, AC; Project administration: AC; Funding acquisition: AXS, YEP, LAQ, ENL, RAV, AC.

## Data Availability

All data produced in the present study are available upon reasonable request to the authors

https://redcovid.uchile.cl/

## ACKNOWLEDGEMENTS

We thank Andrea Ganna at FI, Eric Lander, and Benjamin Neale for facilitating the microarray data to estimate genetic ancestry. Our deepest gratitude to all the participants of the Chilean COVID-19 Biorepository for their generous contribution to the science of Chile. We also thank the technical and professional staff of the Biobank of Tissue and Fluids of the University of Chile for their commitment and support.

## ABBREVIATIONS

COVID-19: Coronavirus disease 19
SARS-CoV-2: Severe acute respiratory syndrome coronavirus-2
GWAS: genome-wide association studies
IC: Informed consent
SOP: Standard Operating Procedures
BTUCH: Biobank of Tissue and Fluids of the University of Chile
CRF: Case Report Form
QC: Quality control
SPREC: Standard Preanalytical Code
SNPs: Single Nucleotide Polymorphisms.

## REFERENCES

1 Hu B, Guo H, Zhou P, et al. Characteristics of SARS-CoV-2 and COVID-19. Nat Rev Microbiol. 2021;19:141–54.

2 Eurosurveillance editorial team. Updated rapid risk assessment from ECDC on coronavirus disease (COVID-19) pandemic in the EU/EEA and the UK: resurgence of cases. Euro Surveill. 2020;25:2008131.

3 Zhou F, Yu T, Du R, et al. Clinical course and risk factors for mortality of adult inpatients with COVID-19 in Wuhan, China: a retrospective cohort study. Lancet (London, England). 2020;395:1054.

4 Mousa M, Vurivi H, Kannout H, et al. Genome-wide association study of hospitalized COVID-19 patients in the United Arab Emirates. EBioMedicine. 2021;74:103695.

5 COVID-19 Host Genetics Initiative. Mapping the human genetic architecture of COVID-19. Nature. 2021;600:472–7.

6 Sirugo G, Williams SM, Tishkoff SA. The Missing Diversity in Human Genetic Studies. Cell. 2019;177:26–31.

7 Petrovski S, Goldstein DB. Unequal representation of genetic variation across ancestry groups creates healthcare inequality in the application of precision medicine. Genome Biol. 2016;17:157.

8 Martin AR, Gignoux CR, Walters RK, et al. Human Demographic History Impacts Genetic Risk Prediction across Diverse Populations. Am J Hum Genet. 2017;100:635–49.

9 Popejoy AB, Fullerton SM. Genomics is failing on diversity. Nature. 2016;538:161– 4.

10 Centers for Disease Control and Prevention (CDC). COVID View Summary ending on July 18, 2020 | CDC. 2020. https://www.cdc.gov/coronavirus/2019-ncov/covid-data/covidview/past-reports/07242020.html (accessed 29 June 2022)

11 Kolin DA, Kulm S, Christos PJ, et al. Clinical, regional, and genetic characteristics of Covid-19 patients from UK Biobank. PLoS One. 2020;15:e0241264.

12 Elliott J, Bodinier B, Whitaker M, et al. COVID-19 mortality in the UK Biobank cohort: revisiting and evaluating risk factors. Eur J Epidemiol. 2021;36:299–309.

13 Raisi-Estabragh Z, McCracken C, Bethell MS, et al. Greater risk of severe COVID-19 in Black, Asian and Minority Ethnic populations is not explained by cardiometabolic, socioeconomic or behavioural factors, or by 25(OH)-vitamin D status: study of 1326 cases from the UK Biobank. J Public Health (Oxf). 2020;42:451–60.

14 UK Biobank. UK Biobank SARS-CoV-2 serology study. 2020.

15 Mikkelsen RB, Gjerris M, Waldemar G, et al. Broad consent for biobanks is best - provided it is also deep. BMC Med Ethics. 2019;20:71.

16 Ministerio de Salud de Chile. Ley 20120 - Sobre la Investigación Científica en el Ser Humano, su genoma y prohibe la clonación humana. 2006. https://www.bcn.cl/leychile/navegar?idNorma=253478

17 Ministerio Secretaria General de la Presidencia de Chile. Ley 19628 - Sobre la Protección de la Vida Privada. 2020. https://www.bcn.cl/leychile/navegar?idNorma=141599

18 World Medical Association (WMA). WMA declaration of Taipei on ethical considerations regarding health databases and biobanks. Second WMA Declar taipei ethical considerations regarding Heal databases biobanks. Published Online First: 2016.

19 Council for International Organizations of Medical Sciences - CIOMS. International ethical guidelines for biomedical research involving human subjects. Bull Med Ethics. 2016;17–23.

20 Parliament and Council of the European Union. Regulation (EU) 2018/1725 of the European Parliament and of the Council of 23 October 2018 on the Protection of Natural Persons with Regard to the 118 Bibliography Processing of Personal Data by the Union Institutions, Bodies, Offices and Agencies and on. Off J Eur Union. 2018.

21 Campbell LD, Astrin JJ, Brody R, et al. BEST PRACTICES: Recommendations for Repositories Fourth Edition. BEST PRACTICES ISBER. 2018. www.isber.org (accessed 22 May 2021)

22 Centers for Disease Control and Prevention (CDC). Biosafety in Microbiological and Biomedical Laboratories (BMBL) 6th Edition | CDC Laboratory Portal | CDC. https://www.cdc.gov/labs/BMBL.html (accessed 30 June 2022)

23 World Health Organization (WHO). Guidance on regulations for the transport of infectious substances 2019–2020: applicable from 1 January 2019. World Health Organization 2019.

24 Altshuler DM, Durbin RM, Abecasis GR, et al. An integrated map of genetic variation from 1,092 human genomes. Nature. 2012;491:56–65.

25 Lindo J, Huerta-Sánchez E, Nakagome S, et al. A time transect of exomes from a Native American population before and after European contact. Nat Commun. 2016;7:13175.

26 De La Fuente C, Galimany J, Kemp BM, et al. Ancient marine hunter-gatherers from Patagonia and Tierra Del Fuego: Diversity and differentiation using uniparentally inherited genetic markers. Am J Phys Anthropol. 2015;158:719–29.

27 Alexander DH, Novembre J, Lange K. Fast model-based estimation of ancestry in unrelated individuals. Genome Res. 2009;19:1655–64.

28 National Institutes of Health (NIH), All of Us Research Program. COVID-19 Participant Experience (COPE). https://databrowser.researchallofus.org/survey/covid-19-participant-experience (accessed 10 November 2022)

29 Pereira AC, Bes TM, Velho M, et al. Genetic risk factors and COVID-19 severity in Brazil: results from BRACOVID study. Hum Mol Genet. 2022;31:3021–31.

30 Verdugo RA, Genova A Di, Herrera L, et al. Development of a small panel of SNPs to infer ancestry in Chileans that distinguishes Aymara and Mapuche components. doi: 10.1186/s40659-020-00284-5

31 Eyheramendy S, Martinez FI, Manevy F, et al. Genetic structure characterization of Chileans reflects historical immigration patterns. Nat Commun. 2015;6. doi: 10.1038/ncomms7472

32 Shelton JF, Shastri AJ, Ye C, et al. Trans-ancestry analysis reveals genetic and nongenetic associations with COVID-19 susceptibility and severity. Nat Genet. 2021;53:801–8.

33 Verma A, Minnier J, Wan ES, et al. A MUC5B Gene Polymorphism, rs35705950-T, Confers Protective Effects Against COVID-19 Hospitalization but Not Severe Disease or Mortality. Am J Respir Crit Care Med. 2022;206:1220–9.

34 Guan W, Ni Z, Hu Y, et al. Clinical Characteristics of Coronavirus Disease 2019 in China. N Engl J Med. 2020;382:1708–20.

35 Fang L, Karakiulakis G, Roth M. Are patients with hypertension and diabetes mellitus at increased risk for COVID-19 infection? Lancet Respir Med. 2020;8:e21.

36 Charlson ME, Pompei P, Ales KL, et al. A new method of classifying prognostic comorbidity in longitudinal studies: development and validation. J Chronic Dis. 1987;40:373–83.

37 Kim Y, Latz CA, DeCarlo CS, et al. Relationship between blood type and outcomes following COVID-19 infection. Semin Vasc Surg. 2021;34:125–31.

38 Need AC, Goldstein DB. Next generation disparities in human genomics: concerns and remedies. Trends Genet. 2009;25:489–94.

39 Bustamante CD, De La Vega FM, Burchard EG. Genomics for the world. Nature. 2011;475:163–5.

40 Landry LG, Ali N, Williams DR, et al. Lack of diversity in genomic databases is a barrier to translating precision medicine research into practice. Health Aff. 2018;37:780–5.

41 Vargas RJ, Cobar OM. The Urgent Need for Management of Biological Samples and Data Accessibility in Latin America. Front Pharmacol. 2021;12. doi: 10.3389/FPHAR.2021.620043

42 Singh S, Cadigan RJ, Moodley K. Challenges to biobanking in LMICs during COVID-19: time to reconceptualise research ethics guidance for pandemics and public health emergencies? J Med Ethics. 2022;48:466–71.

