## Supplementary table 1 for "Cohort Profile of the Chilean COVID-19 Biorepository: a Multicentric initiative for multi-omics research on COVID-19 and LONG-COVID in a Latin American population"

Supplementary Table 1. Prevalence of symptoms reported by donors in the whole biorepository and by severity level.

| | Biorepository | Mild | Hospitalized | Severe hospitalized | Critically ill | Lethal |
|---|---|---|---|---|---|---|
| Fatigue | 1541 (68.1%) | 1355 (79.1%) | 84 (57.5%) | 57 (80.3%) | 44 (29.1%) | 1 (7.7%) |
| Headache | 1521 (67.2%) | 1326 (77.5%) | 82 (56.2%) | 47 (66.2%) | 65 (43.0%) | 1 (7.7%) |
| Myalgia | 1365 (60.3%) | 1157 (67.6%) | 75 (51.4%) | 48 (67.6%) | 83 (55.0%) | 2 (15.4%) |
| Decay | 1322 (58.4%) | 1160 (67.8%) | 73 (50.0%) | 51 (71.8%) | 37 (24.5%) | 1 (7.7%) |
| Anosmia | 1305 (57.7%) | 1193 (69.7%) | 53 (36.3%) | 29 (40.8%) | 29 (19.2%) | 1 (7.7%) |
| Ageusia | 1165 (51.5%) | 1064 (62.1%) | 49 (33.6%) | 28 (39.4%) | 23 (15.2%) | 1 (7.7%) |
| Dyspnea on exertion§ | 1054 (46.6%) | 784 (45.8%) | 98 (67.1%) | 58 (81.7%) | 107 (70.9%) | 7 (53.8%) |
| Persistent dry cough | 988 (43.7%) | 806 (47.1%) | 78 (53.4%) | 38 (53.5%) | 63 (41.7%) | 3 (23.1%) |
| Fever | 968 (42.8%) | 754 (44.0%) | 80 (54.8%) | 48 (67.6%) | 81 (53.6%) | 5 (38.5%) |
| Chills | 951 (42.0%) | 823 (48.1%) | 62 (42.5%) | 41 (57.7%) | 24 (15.9%) | 1 (7.7%) |
| Sore throat | 844 (37.3%) | 752 (43.9%) | 36 (24.7%) | 32 (45.1%) | 24 (15.9%) | 0 (0.0%) |
| Heat and cold sensation | 806 (35.6%) | 708 (41.4%) | 40 (27.4%) | 37 (52.1%) | 21 (13.9%) | 0 (0.0%) |
| Back pain | 801 (35.4%) | 693 (40.5%) | 45 (30.8%) | 32 (45.1%) | 31 (20.5%) | 0 (0.0%) |
| Nasal congestion | 781 (34.5%) | 720 (42.1%) | 23 (15.8%) | 18 (25.4%) | 20 (13.2%) | 0 (0.0%) |
| Diarrhea | 742 (32.8%) | 652 (38.1%) | 39 (26.7%) | 21 (29.6%) | 28 (18.5%) | 2 (15.4%) |
| Chest tightness | 694 (30.7%) | 568 (33.2%) | 54 (37.0%) | 39 (54.9%) | 32 (21.2%) | 1 (7.7%) |
| Difficulty breathing at rest | 678 (30.0%) | 501 (29.3%) | 75 (51.4%) | 51 (71.8%) | 50 (33.1%) | 1 (7.7%) |
| Red eyes | 599 (26.5%) | 533 (31.1%) | 22 (15.1%) | 25 (35.2%) | 19 (12.6%) | 0 (0.0%) |
| Other symptoms | 589 (26.0%) | 438 (25.6%) | 49 (33.6%) | 19 (26.8%) | 81 (53.6%) | 2 (15.4%) |
| Dizziness | 522 (23.1%) | 444 (25.9%) | 34 (23.3%) | 28 (39.4%) | 16 (10.6%) | 0 (0.0%) |
| Numbness | 450 (19.9%) | 363 (21.2%) | 32 (21.9%) | 32 (45.1%) | 23 (15.2%) | 0 (0.0%) |
| Abdominal pain | 440 (19.5%) | 371 (21.7%) | 23 (15.8%) | 25 (35.2%) | 20 (13.2%) | 1 (7.7%) |
| Vomiting | 439 (19.4%) | 379 (22.1%) | 28 (19.2%) | 19 (26.8%) | 13 (8.6%) | 0 (0.0%) |
| Feeling of heavy limbs | 439 (19.4%) | 369 (21.6%) | 24 (16.4%) | 27 (38.0%) | 19 (12.6%) | 0 (0.0%) |
| Sneezing | 397 (17.6%) | 359 (21.0%) | 15 (10.3%) | 15 (21.1%) | 8 (5.3%) | 0 (0.0%) |
| Painful breathing | 386 (17.1%) | 304 (17.8%) | 35 (24.0%) | 30 (42.3%) | 17 (11.3%) | 0 (0.0%) |
| Rhinorrhea | 367 (16.2%) | 324 (18.9%) | 15 (10.3%) | 16 (22.5%) | 12 (7.9%) | 0 (0.0%) |
| New productive cough | 362 (16.0%) | 294 (17.2%) | 25 (17.1%) | 15 (21.1%) | 26 (17.2%) | 2 (15.4%) |
| Inability to move | 341 (15.1%) | 256 (15.0%) | 28 (19.2%) | 34 (47.9%) | 23 (15.2%) | 0 (0.0%) |
| Pneumonia | 283 (12.5%) | 95 (5.5%) | 88 (60.3%) | 56 (78.9%) | 43 (28.5%) | 1 (7.7%) |
| Erythema | 148 (6.5%) | 124 (7.2%) | 12 (8.2%) | 8 (11.3%) | 4 (2.6%) | 0 (0.0%) |
| Eruptions in mouth | 130 (5.7%) | 114 (6.7%) | 8 (5.5%) | 4 (5.6%) | 4 (2.6%) | 0 (0.0%) |
| Skin Rashes | 110 (4.9%) | 86 (5.0%) | 7 (4.8%) | 12 (16.9%) | 5 (3.3%) | 0 (0.0%) |
| Skin discoloration | 68 (3.0%) | 47 (2.7%) | 12 (8.2%) | 2 (2.8%) | 7 (4.6%) | 0 (0.0%) |

§ Sensation of running out of air during physical activity like walking up a flight of stairs
