## Supplementary table 2 for "Cohort Profile of the Chilean COVID-19 Biorepository: a Multicentric initiative for multi-omics research on COVID-19 and LONG-COVID in a Latin American population"

Additional Table 2. Prevalence of reported comorbidities in the whole biorepository and by severity level.

| | **Biorepository** | **Asymptomatic** | **Mild** | **Hospitalized** | **Severe hospitalized** | **Critically ill** | **Lethal** |
|---|---|---|---|---|---|---|---|
| Other chronic diseases | 719 (31.8%) | 38 (22.5%) | 571 (33.4%) | 51 (34.9%) | 24 (33.8%) | 31 (20.5%) | 4 (30.8%) |
| Obesity | 691 (30.5%) | 55 (32.5%) | 529 (30.9%) | 39 (26.7%) | 32 (45.1%) | 35 (23.2%) | 1 (7.7%) |
| Hypertension | 394 (17.4%) | 27 (16%) | 212 (12.4%) | 49 (33.6%) | 26 (36.6%) | 72 (47.7%) | 8 (61.5%) |
| Mental health problems | 233 (10.3%) | 17 (10.1%) | 185 (10.8%) | 14 (9.6%) | 6 (8.5%) | 11 (7.3%) | 0 (0.0%) |
| High cholesterol | 232 (10.3%) | 17 (10.1%) | 161 (9.4%) | 25 (17.1%) | 12 (16.9%) | 17 (11.3%) | 0 (0.0%) |
| Diabetes | 212 (9.4%) | 21 (12.4%) | 103 (6.0%) | 28 (19.2%) | 23 (32.4%) | 35 (23.2%) | 2 (15.4%) |
| Asthma | 148 (6.5%) | 9 (5.3%) | 108 (6.3%) | 13 (8.9%) | 6 (8.5%) | 11 (7.3%) | 1 (7.7%) |
| Cancer (several types) | 98 (4.3%) | 4 (2.4%) | 69 (4.0%) | 15 (10.3%) | 2 (2.8%) | 8 (5.3%) | 0 (0.0%) |
| Anemia | 87 (3.8%) | 2 (1.2%) | 72 (4.2%) | 9 (6.2%) | 2 (2.8%) | 2 (1.3%) | 0 (0.0%) |
| Cardiac problems | 65 (2.9%) | 5 (3.0%) | 37 (2.2%) | 9 (6.2%) | 3 (4.2%) | 8 (5.3%) | 3 (23.1%) |
| Hepatitis | 39 (1.7%) | 1 (0.6%) | 30 (1.8%) | 4 (2.7%) | 3 (4.2%) | 0 (0.0%) | 1 (7.7%) |
| Vascular accident | 33 (1.5%) | 3 (1.8%) | 18 (1.1%) | 6 (4.1%) | 2 (2.8%) | 4 (2.6%) | 0 (0.0%) |
| Weakened immune system | 27 (1.2%) | 1 (0.6%) | 21 (1.2%) | 1 (0.7%) | 1 (1.4%) | 2 (1.3%) | 1 (7.7%) |
| Rheumatoid arthritis | 28 (1.2%) | 3 (1.8%) | 18 (1.1%) | 3 (2.1%) | 2 (2.8%) | 2 (1.3%) | 0 (0.0%) |
| Coronary atherosclerosis | 25 (1.1%) | 0 (0.0%) | 16 (0.9%) | 2 (1.4%) | 1 (1.4%) | 4 (2.6%) | 2 (15.4%) |
| Pulmonary condition | 25 (1.1%) | 0 (0.0%) | 17 (1.0%) | 1 (0.7%) | 1 (1.4%) | 5 (3.3%) | 1 (7.7%) |
| Other rheumatoid diseases | 25 (1.1%) | 1 (0.6%) | 21 (1.2%) | 2 (1.4%) | 0 (0.0%) | 1 (0.7%) | 0 (0.0%) |
| Dialysis | 13 (0.6%) | 0 (0.0%) | 6 (0.4%) | 3 (2.1%) | 0 (0.0%) | 2 (1.3%) | 2 (15.4%) |
| Brain pathology | 12 (0.5%) | 3 (1.8%) | 4 (0.2%) | 1 (0.7%) | 1 (1.4%) | 3 (2.0%) | 0 (0.0%) |
| HIV | 12 (0.5%) | 0 (0.0%) | 7 (0.4%) | 3 (2.1%) | 0 (0.0%) | 1 (0.7%) | 1 (7.7%) |
| Ulcerative colitis or Crohn's disease | 12 (0.5%) | 0 (0.0%) | 9 (0.5%) | 2 (1.4%) | 1 (1.4%) | 0 (0.0%) | 0 (0.0%) |
| Lupus | 7 (0.3%) | 0 (0.0%) | 6 (0.4%) | 0 (0.0%) | 1 (1.4%) | 0 (0.0%) | 0 (0.0%) |
| Pulmonary fibrosis | 5 (0.2%) | 0 (0.0%) | 2 (0.1%) | 1 (0.7%) | 1 (1.4%) | 1 (0.7%) | 0 (0.0%) |
| Tuberculosis | 5 (0.2%) | 2 (1.2%) | 3 (0.2%) | 0 (0.0%) | 0 (0.0%) | 0 (0.0%) | 0 (0.0%) |
| Cystic fibrosis | 2 (0.1%) | 0 (0.0%) | 2 (0.1%) | 0 (0.0%) | 0 (0.0%) | 0 (0.0%) | 0 (0.0%) |
| Transplant of an organ | 2 (0.1%) | 0 (0.0%) | 2 (0.1%) | 0 (0.0%) | 0 (0.0%) | 0 (0.0%) | 0 (0.0%) |
